# Students’ perspectives on communication skills’ course learning modes in two medical schools in Zambia: An interview-based descriptive study

**DOI:** 10.1101/2024.10.02.24314393

**Authors:** Mercy Ijeoma Okwudili Ezeala, John Volk

## Abstract

**Introduction:** This study sought the views of undergraduate medical students on the effectiveness of their modes of instruction in communication skills knowledge and skills acquisition and transfer. Understanding the teaching-learning experiences from the students’ viewpoint could influence communication skills course facilitators to adopt instructional modes and learning experiences that enhance competency development in communication skills. It could also emphasize the need for curriculum developers to review the medical school curriculum, specifying student-centered and active learning pedagogical practices.

**Methods:** The descriptive study involved an in-depth semi-structured interview of eight undergraduate medical students from two medical schools in Zambia, who participated in communication skills training, selected through a purposive nonprobability sampling. The study mostly appropriated Tracy’s phronetic iterative analysis during its thematic analysis of the textual data collected between April and May 2022.

**Results:** The participants affirmed the importance of communication skills to their medical training but, decried the dominance of lecture-based delivery, limited interactive sessions, untimely and inadequate feedback from teachers and peers, inefficacy of teaching communication skills as theory, and limited understanding and distraction from online delivery as factors that hindered communication skills teaching-learning.

**Conclusion:** Although the students expressed communication skills as pivotal to their training and medical practice, their experiences of learning the skills through non-experiential and traditional lecture-based methods did not facilitate the motivation required for mastery and competency, suggesting that facilitators and curriculum planners should reevaluate the teaching-learning modes and process.

## Introduction

The learner remains core to the discourse of learning, notwithstanding the course. Illeris’ learning triangle illustrates the centrality of the learners by situating them in a social, cultural, cognitive, and emotional learning context to acquire meaning [1]. The social dimension positions the learner in a set comprising other people, attitudes, rules, beliefs, and culturally oriented habits; the cognitive addresses knowledge and proficiency, and the emotional enables the learner to react appropriately to external stimuli[1]. Learning becomes the process through which the learner interacts with the social environment and acquires knowledge, emotional stability, and proficiency toward the phenomenon or subject matter [2]. According to Baldwin and Ford [3], the learner’s characteristics, the training design, and the environment might affect the outcome of an educational intervention. Learning experiences that are meaningful and engaging predispose to academic success and effective transfer outside the learning environment [4]. This paper considered the views of selected undergraduate medical students from two medical schools in Zambia on their communication skills learning experiences, focusing on their modes of instruction identified in a study by Ezeala and Volk [5] as one of the areas of concern in teaching-learning communication skills in Zambia undergraduate medical education.

A study among medical students emphasized that communication benefits the patient, healthcare, and the economic system [6]. Communication consists of skills that students can learn. Similarly, Hardee and colleagues [7] identified that intensive communication skills training increased patients’ satisfaction with the medical personnel. The effects of such training lasted even twelve months after the training; hence, training in communication skills is vital in medical education. Although every human being can communicate, relationship-building and maintenance skills sustained by effective communication require attitudes developed and strengthened through meaningful teaching-learning processes. A learning process with methods that are collaborative and experiential and allow medical students to play active roles enhances skills and knowledge transfer and develops self-reflective and independent learning attitudes in the learners [8-10]. Team-based learning, student self-evaluation, facilitator feedback, role-plays, oral presentations, videos, and dramatization were beneficial strategies for learning [11-14]. Likewise, studies that applied interactive learning methods to communication skills facilitated reflective habits and self-directed and team learning [15]. Similarly, Wolff and colleagues [16] discussed techniques that would engage learners in medical education and promote learning rather than retaining didactic teaching delivery that hinders knowledge and skills retention. This evidence suggests that the methods of teaching communication skills implicate the attitude to the mastery of the skills. Lecture-centered learning without modifications to engage students may not effectively enhance competence in communication skills [2].

Despite the suggestions in literature to use learning methods and strategies that involve small groups, collaboration among learners and teachers, simulations, and experiential learning, didactic lectures remain the predominant methods of learning in resource-poor settings with the increased student population and shortage of faculty as experienced currently in many medical schools in Africa. A study in Sudan demonstrated that students in a physiology class found ‘lectures based on problems’ more stimulating and engaging than the conventional lectures despite the large class population [17]. Another study indicated that the team-based lecture method facilitated interaction, self-reflection, and deep learning, unlike traditional lectures [10]. These studies, therefore, imply that conventional lectures are less effective modes of teaching in medical education; however, a teacher can modify the lectures to make the learning interactive, engaging, and meaningful.

Based on the study by Ezeala and Volk [2] involving communication skills training in undergraduate medical education at the University of Zambia (UNZA) and Mulungushi University (MU), which highlighted that sixty-eight percent of the participants reported didactic lectures as their main training mode, this study sought the views of students from these two universities toward their teaching-learning methods in communication skills. The intent was to understand from the learners how to make communication skills learning meaningful for skills and knowledge transfer and acquisition by gaining their perception regarding the effectiveness of the instruction modes.

## Materials and methods

### Settings

Two of the eight medical schools in Zambia participated in this study and their geographical locations were in different provinces. The School of Medicine, University of Zambia (UNZA), Ridgeway Campus, is in Lusaka and has offices at Ridgeway and the University Teaching Hospital. Its’ Bachelor of Medicine and Bachelor of Surgery degree (MBChB), which is the focus of this study, offered communication skills to the clinical students in their fifth year as a component of a course combining communication skills, healthcare ethics, and professionalism. Mulungushi University (MU) has its medical school domiciled in Livingstone, Southern Province, and offered communication skills to second-year preclinical students in combination with psychology under behavioral sciences during this study period. Despite the difference in the academic years of the participants, the two medical schools observe competency-based curricula with ‘similar contents’ [5]. Communication skills content in both schools includes topics such as history-taking, communicating with different patients, breaking bad news, difficult situations, intercultural communication, gender roles in communication, collaborative practice in healthcare settings, and healthcare documents.

### Study design and ethical considerations

Data collection for this study occurred between 22 April and 27 May 2022. The interview venues varied among the two medical schools that participated in the study and were chosen to ensure the participants’ comfort and convenience, and private for discussions. Eight participants purposively selected through maximum variation based on study level and gender took part in the study, stratified into two females and two males, respectively, from the participating institutions, aged between 20 and 25 years. The participants were registered for and offered the communication skills course and participated in the study by Ezeala and Volk [2]. Additionally, the study did not seek further participants because the data collected from the eight participants were exhaustive, yielding detailed information and comprehensive understudying of the student’s perception of their communication skills’ teaching-learning approaches.

The study obtained ethical approval from the ethical review committee of MU, a clearance from the National Health Research Authority (NHRA), and permission from the deans of the two medical schools. Before the interviews, the researchers sent an electronic copy of the information and consent forms explaining the purpose of the interviews, inviting the participants to indicate their willingness to participate by identifying the appropriate time. The investigators conducted the interviews personally to understand the needed information clearly. The researchers presented the participants with a hard copy of the interview consent form for their attestation after ascertaining the participants’ understanding of the purpose of the interviews and sought permission to audiotape the proceedings. The researchers stored the consent forms securely. The interview guide (S1 Appendix) helped structure the proceedings but did not hinder the flexibility and flow of information. The discussions lasted between 15 and 35 minutes each. The researcher conducted an initial verbatim transcription of the audiotapes shortly after the interviews to ensure retention and accurate recording before typing and filing the transcripts. The transcripts have no personally identifiable information as the researchers anonymized the data to maximize the participants’ confidentiality by preserving their privacy. The participants’ identifiers were not related to the participants and were only meaningful to the researchers.

### Data analysis

While many researchers affirm that data analysis in qualitative research has no recipe that must be strictly adhered to for a particular result [18-20], many qualitative methodologies, such as grounded theory and phenomenology, have defined practical analytical processes to ensure credibility and dependability in the results. This study adopted Tracy’s phronetic iterative analysis [21] in its thematic analysis of the textual data to help guide and provide a degree of thoroughness and explicitness in describing how this study arrived at its themes, actively and consciously. The phronetic iterative analysis focuses on parts of the collected data that manage pragmatic issues, rather than on the entire data and bases its categorization on the participants’ narratives [21]. Its emphasis on the context of research and the effects of the analysis on the practical lives of the participants facilitate a concentrated effort in addressing a study’s objectives; however, the choice does not imply that this study used the analytic procedure as a cookbook. This approach to qualitative analysis provided the order this study needed to maintain quality while allowing it the flexibility to respond to its exigencies.

The descriptive first level, analytic second level coding, and developing a codebook compose the three stages of the phronetic iterative analysis. The analytic procedures included manual and computer-assisted output using Microsoft Word documents. During the organization and data preparation stage, the researchers prepared a face sheet containing information about each interviewee comprising the identification marker, gender, study level, interview date, venue, and the main interview question. The audio record of the discussions was transcribed and stored as a Microsoft Word document. After organizing the data, the investigators read and reread the transcribed data, noting reflections about the data in a notebook and underlined words, phrases, or sentences that resonated with the study objectives from the printed transcribed texts. The iterative reading and interactions with the scripts provided an intimate understanding of the participants’ views, what qualitative researchers call immersion [22, 20]. At the first-level or primary-cycle coding, the researchers read through segments of the transcriptions and assigned words or phrases that evoked or reflected the meanings the texts harbored. They wrote these in red color at the edge of the Word document while underlining significant verbatim statements from the participant. S2 Appendix is an example of the primary circle coding.

During the second level, secondary-cycle coding, the researchers consulted the literature on constructivist learning and empirical studies concerning the most appropriate mode of delivering communication skills to learners, selected and combined the significant codes from the first level coding, thereby condensing or “zooming” the codes to “focused codes” [20] or “themes” [23] that are analytical and reflective of empirical themes in communication skills course delivery. Related codes occupied the same category with an umbrella theme. The secondary-cycle coding provided an interpretive ‘working skeleton’ [21] that led to a codebook defining themes understood by this study and excerpts from the participants’ narratives that highlighted them. The codebook used in this study refers to the notes organized under the themes or codes reflecting the needs or problems highlighted from the data and literature, the study’s assigned meaning, and examples from the transcripts. The researchers kept writing reflections about the data and the analytic procedures throughout the analysis, including notes that deviated from the developing narratives.

## Results

### Description of the participants

The participants comprised 2 male and female students from the two participating institutions, pooled from years five and two. The age of the participants ranged from 20 to 25. Tables 1 and 2 summarize the information relating to the participants and the interview. The participants ‘identifiers in Table 2 consist of numbers and alphabets meaningful only to the researchers.

**Table 1.**
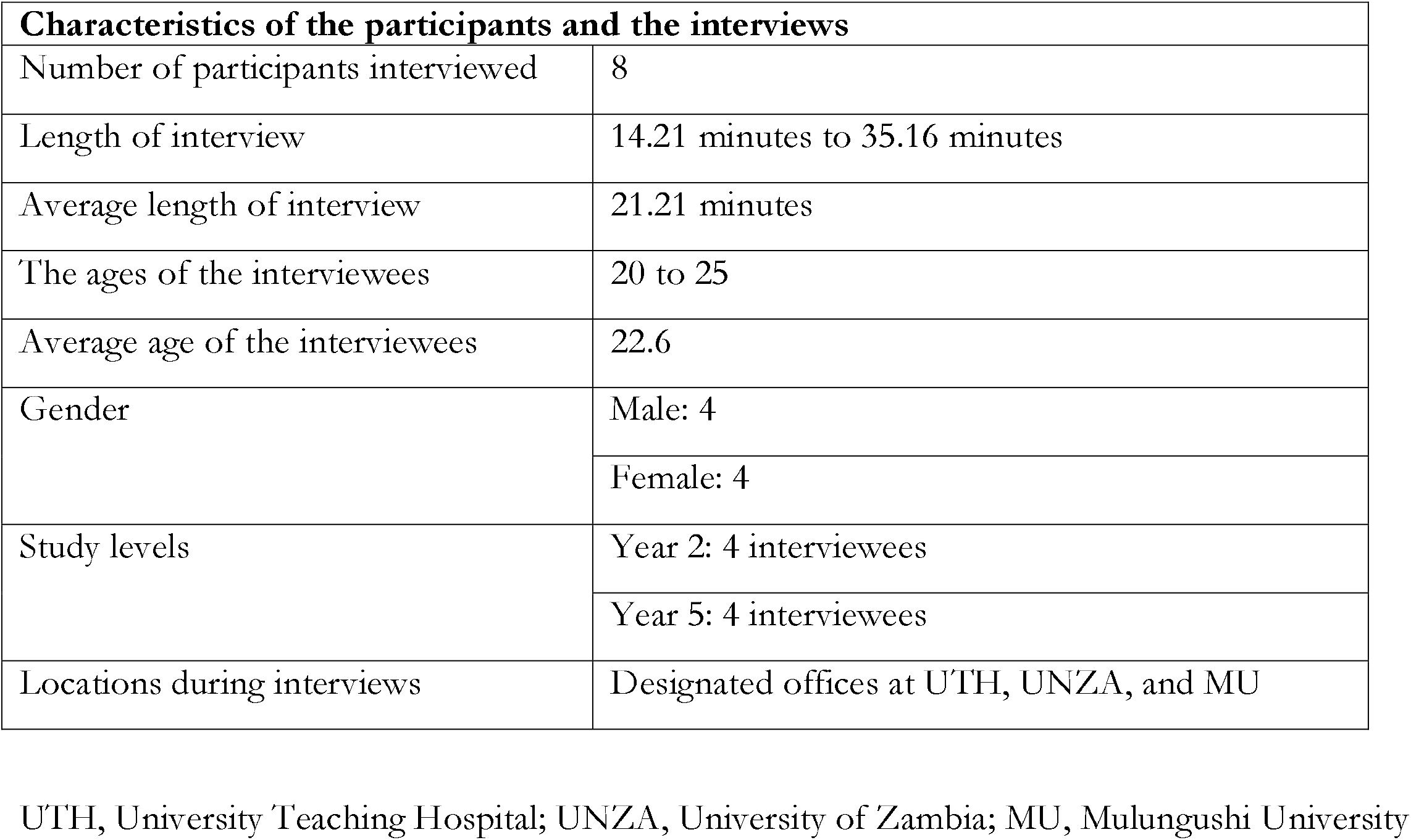
Participants and interview details.

**Table 2.** Additional data on the interviewees.

| Participant code |  |
| --- | --- |
|  | ME1 |
|  | MT1 |
|  | US1 |
|  | UK1 |
|  | MR11 |
|  | MH11 |
|  | KU11 |
|  | UE11 |
| **Gender** | Male: 4<br>Female: 4 |
| **Age: years** | 20-25 |
| **Study level** | Years 2 and 5 |

### Study mode and subthemes

Table 3 captures the theme of the study mode and its subthemes arising from the participants’ views.

**Table 3.** Themes and subthemes.

| Theme | Subthemes |
| --- | --- |
| Modes of learning<br>communication skills | Preponderance of lectures |
|  | Insufficient interactive sessions and non-interactive lectures |
|  | Teaching it as a theory rather than practically |
|  | Untimely and little feedback from teachers and peers |
|  | Online delivery was challenging and distracting |

### Preponderance of lectures and theory

While almost all the interviewees found communication skills interesting because:

> *‘… most of these other courses that we do require us to just sit and listen, but then communication skills course was more interactive, given we get to interact with the lecturers as well*…’ (MR11)

> *Communication skills in the practice of medicine … allowed myself as a student and I assume, other students, to observe and reflect on the variation between classroom teaching and real-life practice in this field. The integration of communication skills into medicine bridged the gap between the reality in this field of practice and the real environment of classroom teaching, which is the main cause of anxiety for a student coming from the preclinical years*. (UK1)

They still complained about the predominantly lecture-based teaching mode and the need to include more interactive teaching approach such as role-plays, team discussions, and simulated learning. One of the participants stated:

> *‘I think the way we were trained could be better improved. The way the information was delivered to us could be better. … When you lecture or teach a certain group, you are not talking to everyone …. …, most of the times we have our lectures we’re basically just receiving, we’re not actually reciprocating*.*’* (MT1)

MH11 reminisced about the video she saw during her communication skills lectures and asserted that she could still picture the events and that videos should be added to communication skills lectures. Another interviewee delineated the practicality of communication skills and the limitations of teaching them theoretically. According to the participant:

> *I consider communication skills to be practical instead of theory …. I think it would help if it were to be put as a practical course instead of sitting and waiting to be taught…. For you to teach it as practical, you’ll want to observe your students … in the ward to see how they best interact with their patients* (ME1).

KU11 also complained about the lack of practice during their learning experiences and highlighted the need for examining communication skills practically:

> *…just not practicing enough, everything was just theoretical; so, the chances of people carrying that knowledge are slim. …more practice; practice sessions and also practical exams because when it’s just hearing or just writing to pass, but practical exams can make it stick more*. (KU11)

### Limited interactive sessions and poor feedback

Further to these statements, another interviewee specified the need for participatory learning through role-plays and timely feedback rather than lectures:

> *Apart from lecturing, we could set up a scenario-based type of learning, where we could play out roles or maybe a specific kind of case scenario where specific students are chosen to play so that they know how to interact because a lot of us really don’t know how it feels in a real situation; most of us come here without experience. So, it’s different for us when we enter the field; … it’s better if we were given those scenarios when teaching* (MT1).

> *So, I feel, maybe introducing, I don’t know if I can call it drama or sketches, like where students may, maybe … role play as the patient, then they’re subdivided like that, … then people will present and then from there now, their friends, or maybe the entire class or the moderator will say, “There is a mistake you did and such and such*.*” Then they can be role-playing things, like breaking bad news, and then because it can be so easy to memorize how to break down bad news; but when somebody is given a role, I think even for that person involved, as well as the people watching, I think it sticks more because they’ll always remember, ‘oh, that’s the mistake that my friend made there’* (US1).

A participant also commented about the dissatisfaction with lecturers who do not facilitate positive relationships with the students during communication skills learning:

> *‘… It’s an interactive course and it has to be a person that’s interactive with the students…. Because I feel the more students are able to interact, the more they are able to ask questions. I think the real thing will be an interactive and accommodating facilitator*.*’* (US1).

### Problems with online learning

Participants from the UNZA had most of their communication skills classes online because of COVID-19. The interviewees expressed dissatisfaction with the online mode of delivering communication skills because of distractions. According to an interviewee:

> *…the online one disadvantaged me personally because level of concentration … would be different, not interactive. … I’ll have other things to do… but in the end, you’ll find that there’s really nothing you’ve gotten from the lecture … you’ll have to go and study on your own, which sometimes would be hard because some things which were explained by the lecturer, you’ve missed them. So, for me, the online one was bad, but I feel if it was done physically, it was going to be more helpful, too, and I would understand more than I did during the online lesson* (ME11).

> *… I learned my communication skills during the pandemic and in most cases, our lectures were done online and the time we had to have the experience with our perfector wasn’t that much. …; I think that’s the most challenge I encountered* (UK1).

KU11 also complained about the online nature of their communication skills delivery.

> *We learned a lot, but the way it was online … didn’t help me a lot*.*’* (KU11)

The participants expressed positive attitudes toward learning communication skills. They viewed the course as central to medical training and practice; however, their expressed concerns about aspects of their predominantly lecture-based teaching-learning mode is an issue of concern.

## Discussion

This study explored the perception of undergraduate medical students at MU and the UNZA regarding their predominantly lecture-based communication skills teaching-learning mode, concentrating on the subthemes relating to ineffective teaching methods. Other qualitative studies also applied thematic analysis to identify the barriers to teaching communication skills in Spanish medical schools and South Africa [6, 24]. Qualitative studies in communication skills have yielded much information in the field [25-26].

The participants all affirmed that the communication skills course is vital to undergraduate medical education because of its central role in relationship-building and enhancing understanding. Several studies also demonstrated that undergraduate medical students consider communication skills crucial in building the confidence of medical students for effective interactions [27-29]. Although this current study’s participants lauded the importance of the communication skills training they received, they still expressed dissatisfaction with aspects of their training relating to the mode of course delivery.

Methods for delivering communication skills effectively to medical students have been the context of several studies. Agago and colleagues [12] posited that undergraduate medical students preferred interactive teaching-learning modes and demonstrated that a simulated patient-based approach enhanced learning and performance in communication skills more than the case-based technique in a medical school in Ethiopia. Another study in Sudan compared two teaching modes, verifying that the students enjoyed their sessions in ‘lectures based on problems,’ participated actively, and retained information on physiology more than in the traditional lecture approach [17]. A study reported an improvement in the interview skills of third-year medical students who experienced standardized patient training in the group trained with conventional lectures in a randomized controlled trial [30]. Similarly, a randomized crossover study conducted in a medical school in Saudi Arabia demonstrated students’ preference for team-based learning and better performance in clinical reasoning than in lectures [31]. These highlighted studies align with the participants’ views that traditional lectures in communication skills are not as effective in enhancing communication skills competencies as participatory teaching-learning modes such as modeling appropriate communication skills before learners at the clinics [32] and using role-plays, simulated patients, and experiential learning [33-35]. Despite the students’ partiality to interactive and small group learning and their effectiveness in skills acquisition, the cost and time implications of using these interactive strategies affect the adoption of communication skills, especially in settings with a high student population and limited resources. Concurring with this view, Chege and Njengere [36] posit that lecture-based training might serve well in resource-limited settings, but it is not the best for skills transfer. It is not, therefore, farfetched to declare that the teaching-learning approach that will improve communication skills in medical schools like MU and the UNZA must consider the learning context and economical yet efficient mode of teaching-learning.

The clinical students in the interviews complained about their difficulties with online learning as a teaching mode. Several studies reported the challenges that medical students, who were not used to online learning platforms, faced during the COVID-19 pandemic [37-38]; others identified the positive aspects of online learning, especially when blended with face-to-face mode for learning clinical skills [39-41]. Like this current study that concentrated on communication skills in medical education, Lange and colleagues [42] investigated the viability of online history taking, comparing the students’ evaluation of their learning through the online and lecture training modes. Though the students performed well in the online history taking, which they conducted as teams, and commended the flexibility it afforded them, they still preferred the traditional lectures in history taking. This preference could be because the students would still prefer physical lectures to online learning due to the versatility and interactions that face-to-face teaching-learning affords.

The participants from this study considered communication skills a practical course, which implies that using online mode to teach communication skills requires careful planning of the teaching-learning activities to enable effective interactions among the teachers and learners. Coincidentally, research suggests that communication skills learned in the classrooms rarely translate to medical practice [43-44] because of the disconnect between content and theory learned in the class and the practice of communication at the clinics [45]. Furthermore, an empirical study [46] identified that many medical schools rarely meet the guidelines and recommendations for teaching communication skills because they teach communication skills alongside theoretically-based subjects such as ethics and psychology independent of the clinical courses. Students had minimal opportunities to practice and the training limited the assessments to the traditional written examinations without structuring them in simulated or hospital settings. The practicality of communication skills further requires feedback that immediately addresses communication behavior, hence the need for interactive approaches to teaching-learning.

## Conclusion

Undergraduate medical students that participated in this study considered learning communication skills appropriate for developing communicative competence in clinical and interpersonal skills; however, the preponderance of traditional lectures, poor interactive sessions and feedback, and improper curriculum representation and administration of the communication skills course were factors the participants considered ineffectual teaching mode. Different teaching modes including lectures, team-based learning, role-plays, presentations, and simulations should be combined during communication skills teaching-learning sessions. Communication skills facilitators must proactively apply pedagogical modes that enable skills acquisition and transfer. This will enhance the establishment of communication skills as an important aspect of medical education.

## Data Availability

All relevant data are within the manuscript and its Supporting Information files

## Supporting information

1. S1 appendix. The interview guide
2. S2 appendix. The primary circle coding

## References

1. Pätzold H. Learning and teaching in adult education: Contemporary theories. 1st ed. Leverkusen Opladen: Barbara Budrich Publishers; 2011.

2. Ezeala MIO, Volk J. A survey of students’ attitudes and comments about needed changes in communication skills learning in Zambia: A descriptive study. IJIRME. 2023; 2(4): 165–171. 10.58806/ijirme.2023.v2i4n06.

3. Baldwin TT, Ford JK. Transfer of training: a review and directions for future research. Personnel Psychology. 1988; 41(1): pp. 63–105. 10.1111/j.1744-6570.1988.tb00632.x.

4. Evans C, Mujis D, Tomlinson D. Engaged student learning: High impact strategies to enhance student achievement. York, GB: Higher Education Academy; 2015.

5. Ezeala MIO, Volk J. Relationships between undergraduate medical students’ attitudes toward communication skills learning and demographics in Zambia: A survey-based descriptive study. J Educ Eval Health Prof. 2023; 20.16. 10.3352/jeehp.2023.20.16.

6. Ruiz-Moral R, García de Leonardo C, Cerro Pérez A, Caballero-Martínez F, Monge-Martín D. Barriers to teaching communication skills in Spanish medical schools: A qualitative study with academic leaders. BMC Med Educ. 2020; 20(41). Available from: 10.1186/s12909-020-1944-9.

7. Hardee JT, Rehring TF, Cassara JE, Weiss K, Perrine N. Effect and durability of an in-depth training course on physician communication skills. Perm J. 2019; 23(18-154). Available from: 10.7812/TPP/18-154.

8. Zeng H, Chen D, Li Q, Wang X. Effects of seminar teaching method versus lecture-based learning in medical education: A meta-analysis of randomized controlled trials. Med Teach. 2020; 42(12):1343–1349. 10.1080/0142159X.2020.1805100.

9. Mata A, de Azevedo KPM, Braga LP, de Medeiros GCBS, Segundo VHdO, Bezerra INM, et al. Training in communication skills for self-efficacy of health professionals: A systematic review. Hum Resour Health. 2021; 19(30). Available from: 10.1186/s12960-021-00574-3.

10. Salih KEMA, El-Samani EFZ, Bilal JA, Hamid EK, Elfaki OA, Idris MEA, et al. Team-based learning and lecture-based learning: Comparison of Sudanese medical students’ performance. Adv Med Educ Pract. 2021; 12(24 December):1513–1519. 10.2147/AMEP.S331296.

11. Nair B. Role play - an effective tool to teach communication skills in pediatrics to medical undergraduates. J Educ and Health Promot. 2019; 8(18). Available from: 10.4103/jehp.jehp_162_18.

12. Agago T, Wonde S, Bramo S, Asaminew T. Simulated patient-based communication skills training for undergraduate medical students at a university in Ethiopia. Adv Med Educ Pract. 2021; 12(25 June):713–721. 0.2147/AMEP.S308102.

13. Pless A, Hari R, Brem B, Woermamm U, Schnabel KP. Using self and peer video annotations of simulated patient encounters in communication training to facilitate the reflection of communication skills: An implementation study. GMS J Med Educ. 2021; 38(3), Doc55. Available from: 10.3205/zma001451.

14. Qaiser R, Fadden P, Rustom S, Shaw J. Real-time peer-to-peer observation and feedback lead to improvement in oral presentation skills. Cureus, 2022; 14(2), p. e21992. Available from: 10.7759/cureus.21992.

15. Tan XH, Foo MA, Lim SLH, Lim MBXY, Chin AMC, Zhou J, et al. Teaching and assessing communication skills in the postgraduate medical setting: A systematic scoping review. BMC Med Educ. 2021; 21(1). Available from: 10.1186/s12909-021-02892-5.

16. Wolff M, Wagner MJ, Poznanski S, Schiller J. Santen S. Not another boring lecture: Engaging learners with active learning techniques. J Emerg Med. 2015; 48(1):85–93. 10.1016/j.jemermed.2014.09.010.

17. Alaagib N, Musa O, Saeed A. Comparison of the effectiveness of lectures based on problems and traditional lectures in physiology teaching in Sudan. BMC Med Educ. 2019; 19(1). Available from: 10.1186/s12909-019-1799-0.

18. Guest G, MacQueen K, Namey E. Applied thematic analysis. Thousand Oaks: SAGE Publications, Inc; 2012.

19. Humble Á, Radina M. Introduction: real stories of how this volume happened. In: Humble A, Radina M, editors. How qualitative data analysis happens: Moving beyond “themes emerged”. New York: Routledge; 2019. pp. xix-xxix.

20. Järvinen M, Mik-Meyer N. Analysing qualitative data in social science. In: Järvinen M, Mik-Meyer N, editors. Qualitative analysis: Eight approaches for the social sciences. London: SAGE Publications Ltd; 2020. pp. 1–27.

21. Tracy S. Qualitative research methods: Collecting evidence, crafting analysis, communicating impact. 2nd ed. Hoboken: John Wiley and Sons; 2020.

22. Grbich C. Qualitative data analysis: An introduction. 2nd ed. London: SAGE Publications Ltd; 2013.

23. Saldaña J. The coding manual for qualitative researchers. 3rd ed. Thousand Oaks, CA: SAGE; 2016.

24. Ganca L, Gwyther L, Harding R, Meiring M. What are the communication skills and needs of doctors when communicating a poor prognosis to patients and their families? A qualitative study from South Africa. S Afr Med J. 2016; 109(9):940–944. 10.7196/SAMJ.2016.v106i9.10568.

25. Perron JN, Cronauer KC, Hautz SC, Schnabel KP, Breckwoldt J, Monti M, et al. How do Swiss medical schools prepare their students to become good communicators in their future professional careers: A questionnaire and interview study involving medical graduates, teachers and curriculum coordinators. BMC Med Educ. 2018; 18(285). Available from: 10.1186/s12909-018-1376-y.

26. Graf J, Zipfel S, Wosnik A, Mohr D, Herrmann-Werner A. Communication skills of medical students: survey of self- and external perception in a longitudinally based trend study. BMC Med Educ. 2020; 20(149). Available from: 10.1186/s12909-020-02049-w.

27. Nayak R, Kadeangadi D. Effect of teaching communication skills to medical undergraduate students: An exploratory study. Indian Journal of Community and Family Medicine. 2019; 5(2):108–113. 10.4103/IJCFM.IJCFM_66_19.

28. Jeddi F, Ghaffary F, Farrahi R. The relationship between communication skills and intellectual health in senior-year students of paramedicine school at Kashan University of Medical Sciences 2019. The Open Public Health Journal. 2020; 13(1):484–488. 10.2174/1874944502013010484.

29. Mangan J, Rae J, Anderson J, Jones D. Undergraduate paramedic students and interpersonal communication development: A scoping review. Adv Health Sci Educ Theory Pract. 2022; 27(4):1113–1138. 10.1007/s10459-022-10134-6.

30. Geoffroy PA, Delyon J, Strullu M, Dinh AT, Duboc H, Zafrani L, et al. Standardized patients or conventional lecture for teaching communication skills to undergraduate: A randomized controlled trial. Psychiatry Investig. 2022; 17(4):299–305. 10.30773/pi.2019.0258.

31. Imran M, Halawa TF, Baig M, Almanjoumi AM, Badri MM, Alghamdi WA. Team-based learning versus interactive lecture in achieving learning outcomes and improving clinical reasoning skills: A randomized crossover study. BMC Med Educ. 2022; 22(1): 348. Available from: 10.1186/s12909-022-03411-w.

32. Taveira-Gomes I, Mota-Cardoso R, Figueiredo-Braga M. Communication skills in medical students - an exploratory study before and after clerkships. Porto Biomed J. 2016; 1(5):173. 10.1016/j.pbj.2016.08.002.

33. Koponen J, Pyörälä E, Isotalus P. Communication skills for medical students: Results from three experiential methods. Simulation & Gaming. 2014; 45(2): 235–254. 10.1177/1046878114538915.

34. Ferreira-Padilla G, Ferrández-Antón T, Baleriola-Júlvez J, Braš M, Đorđević V. Communication skills in medicine: Where do we come from and where are we going? Croat Med J. 2015; 56(3): 311–314. 10.3325/cmj.2015.56.311.

35. Burgess A, van Diggele C, Roberts C, Mellis C. (2020) Team-based learning: Design, facilitation and participation. BMC Med Educ. 2020; 20(Suppl 2): 461. Available from: 10.1186/s12909-020-02287-y.

36. Chege M, Njengere D. The state of academic writing in Kenyan universities: Making a case for Kenyan universities to re-conceptualize their approach to teaching academic writing. Journal for Research and Practice in College Teaching. 2018; 3(1): 10–46.

37. Al-Balas M, Al-Balas HI, Jaber HM, Obeidat K, Al-Balas H, Aborajooh EA, et al. Distance learning in clinical medical education amid COVID-19 pandemic in Jordan: Current situation, challenges, and perspectives. BMC Med Educ. 2020; 20(1): 341. Available from: 10.1186/s12909-020-02257-4.

38. Baticulon RE, Sy JJ, Alberto NRI, Baron MBC, Mabulay REC, Rizada LGT, et al. Barriers to online learning in the time of COVID-19: A national survey of medical students in the Philippines. Med. Sci. Educ. 2021; 31(2): 615–626. 10.1007/s40670-021-01231-z.

39. Rajab M, Gazal A, Alkattan K. Challenges to online medical education during the COVID-19 pandemic. Cureus. 2020; 12 (7): e8966. Available from: 10.7759/cureus.8966.

40. AlQhtani A, AlSwedan N, Almulhim A, Aladwan R, Alessa Y, AlQhtani K. Online versus classroom teaching for medical students during COVID-19: Measuring effectiveness and satisfaction. BMC Med Educ. 2021; 21(1): 452. (Online) Available from: 10.1186/s12909-021-02888-1.

41. Tayem YI, Almarabheh AJ, Abo Hamza E, Deifalla A. Perceptions of medical students on distance learning during the COVID-19 pandemic: A cross-sectional study from Bahrain. Adv Med Educ Pract. 2022; 13(12 April): 345–354. 10.2147/AMEP.S357335.

42. Lange S, Krüger N, Warm M, Op den Winkel M, Buechel J, Hubert J, et al. Online medical history taking course: Opportunities and limitations in comparison to traditional bedside teaching. GMS J Med Educ. 2022; 39(3): Doc34. Available from: 10.3205/zma001555.

43. Rosenbaum ME, Axelson R. Curricular disconnects in learning communication skills: What and how students learn about communication during clinical clerkships. Patient Educ Couns. 2013; 9(1): 85–90. 10.1016/j.pec.2012.10.011.

44. Møller JE, Kjaer LB, Helledie E, Nielsen LF, Malling BV. Transfer of communication teaching skills from university to the clinical workplace - does it happen? A mixed methods study. BMC Med Educ. 2021; 21(1): 433. Available from: 10.1186/s12909-021-02834-1.

45. Yardley S, Irvine AW, Lefroy J. Minding the gap between communication skills simulation and authentic experience. Med Educ. 2013; 47(5): 495–510. 10.1111/medu.12146.

46. Ruiz-Moral R, Garcia de Leonardo C, Caballero-Martínez F, Monge-Martín D. Medical students’ attitudes toward communication skills learning: comparison between two groups with and without training. Adv Med Educ Pract. 2019; 10(Feb.12): 55–61. 10.2147/amep.s182879.

